# Transcriptomic Immune-related Signature Predictive of Chemoradiotherapy Response in Anal Squamous Cell Carcinoma

**DOI:** 10.64898/2026.03.12.26348072

**Authors:** Soledad Iseas, Mariano Golubicki, Ezequiel Lacunza, Diego Prost, Sarah Bouchereau, Chloe Lahaie, Nabil Baba-Hamed, Eric Raymond, Julien Adam, Martin Carlos Abba

## Abstract

Anal squamous cell carcinoma (ASCC) is a rare malignancy associated with high-risk HPV, with rising incidence among younger adults. While immunotherapy has improved outcomes in metastatic ASCC, treatment for localized disease remains largely unchanged, with high recurrence rates. This study provides comprehensive exome and transcriptome profiling of 40 stage I-III non-metastatic ASCC patients treated with curative chemoradiotherapy (CRT) to identify predictors of treatment response and progression-free survival. Transcriptomic analysis revealed 350 differentially expressed genes between complete responders (CR) and non-complete responders (NCR) (p-value<0.01; FC>2). CR was associated with modulation of immune-related pathways, cytokine production, epidermis development, cell differentiation, and signaling pathways associated with TNFA/NFkB and epithelial to mesenchymal transition. Immune infiltrate analysis showed significant enrichment of CD8+ central memory T cells (p=0.008) in CR cases, correlating with increased tertiary lymphoid structure and improved overall (p=0.0026) and disease-free survival (p=0.0098). Exome-seq identified alterations in novel and known cancer driver genes without association to CRT response, despite high tumor mutational burden (TMB) was significantly associated with shorter overall (p=0.03) and disease-free survival (p=0.027) compared with low TMB cases. These findings highlight the potential of incorporating gene expression signatures (e.g., *FDCSP, ALDOB, ADGRB1, SPINK7*) alongside immune-related markers into clinical practice to enhance the prediction of treatment response and guide personalized therapies in ASCC. A robust and functionally active immune microenvironment, characterized by specific T and B cell populations and the presence of tertiary lymphoid structures, emerges as a hallmark of complete response and improved survival in ASCC patients undergoing chemoradiotherapy.

## 1. INTRODUCTION

Anal squamous cell carcinoma (ASCC) is a rare but increasingly common malignancy strongly associated with high-risk human papillomavirus (HPV) infections that have a higher incidence among younger adults lately (1,2) Strategies such as HPV vaccination and screening in high-risk populations are being adopted to mitigate this trend, emphasizing the impact of these preventive measures (3,4). Additional risk factors, though less strongly associated, include HIV infection, tobacco use, immunosuppression, and autoimmune conditions such as Crohn’s disease, which may contribute through alternative pathogenic mechanisms (5,6).

Recently, new therapeutic alternatives based on immunotherapy have expanded the treatment landscape. The combination of carboplatin-paclitaxel with retifanlimab has shown superiority over chemotherapy alone in first-line treatment for metastatic or recurrent ASCC, considered as a new first-line standard (7). Before this, promising results from early-phase trials of nivolumab and pembrolizumab in chemotherapy-refractory patients have reported objective responses in 11-24% of cases, suggesting new therapeutic possibilities and diversifying treatment strategies (8–11). While novel combinations of chemotherapy, immunotherapy and radiotherapy are ongoing being explored (12), the standard treatment for the curative setting has remained largely unchanged since the 1990s, and recurrence rates remain high (25-40%) particularly in locally advanced stages (12,13).

Efforts to elucidate the molecular and immune mechanisms driving HPV-associated ASCC are continuously emerging. However, current evidence predominantly stems from retrospective studies often lacking associated clinical and treatment parameters. Despite these limitations, significant advances have been hypothetically outlined across multiple omics levels. Genomic and epigenomic alterations, including *PIK3CA* mutations and DNA methylation patterns, have emerged as prognostic and predictive biomarkers, guiding targeted therapies and enabling disease monitoring (14)(15). On the other hand, the tumor microenvironment in HPV-associated ASCC plays a pivotal role in immune evasion through the actions of Tregs and myeloid-derived suppressor cells (MDSCs), which suppress immune responses and promote T-cell exhaustion. Immune-related signatures show promise in enhancing the efficacy of immune checkpoint blockade therapies (15)(16). HPV-driven ASCC fosters an immunosuppressive microenvironment through regulatory T cells (Tregs) and PD-L1 expression, hindering antitumor immunity (16)(17). Collaborative research remains crucial to advancing personalized therapies and improving patient outcomes in this rare malignancy.

In this study, we analyzed a cohort of NM-ASCC patients to identify mutational, transcriptomic, and immune-based biomarkers associated with chemoradiotherapy outcomes. Our goal was to gain a deeper molecular understanding of the mechanisms underlying this rare malignancy and to uncover prognostic and predictive markers of individual treatment response.

## 2. MATERIALS & METHODS

### 2.1 Anal cancer cohort

This retrospective study comprised 97 consecutive eligible non-metastatic anal cancer patients recruited between 2010 and 2017 who were treated with curative intent at the oncology unit of Hospital Paris Saint Joseph (HPSJ) in Paris, France. The protocol was approved by the ethics committee of the HPSJ institution. Patients had to provide informed consent for their data collection according to the recommendation of the ethics committee and in accordance with the European Union General Data Protection Regulation (GDPR).

Inclusion criteria were at least 18 years old, available pretreatment formalin-fixed paraffin-embedded (FFPE) biopsy, histologically confirmed squamous cell carcinoma, clinical stage cTNM I-III disease, and completed definitive CRT as their primary therapeutic approach. Patients with anal adenocarcinoma, in situ squamous cell carcinoma or other histological cancer subtypes were excluded. Clinical and pathologic data were retrieved from the electronic charts. Initial clinical staging was based on anoscopy and digital anal examination, thorax–abdomen computed tomography (CT) scan, pelvic magnetic resonance imaging (MRI), and FDG-PET/TC.

### 2.2 Treatment and Follow-up

Radiotherapy was performed using either 3D conformal or intensity-modulated techniques (IMRT), with a median dose of 54 Gy in 30 daily fractions over 5.5 weeks. The three chemotherapy regimens delivered concomitantly with radiotherapy were: 1) mitomycin 12 mg/m2 IV bolus, day 1-29 (maximum dose 20 mg) with 5-FU 1,000 mg/m2 on days 1–4 and 29–32 by continuous 24-h IV infusion; 2) mitomycin 12 mg/m2 IV bolus, only day 1 (maximum dose 20 mg) with capecitabine 825 mg/m2 twice daily on each radiotherapy treatment day, and 3) cisplatin 60 mg/m2 on days 1 and 29, with 5-FU 1000 mg/m2 on days 1–4 and 29–32 by continuous 24-h IV infusion. All HIV-positive patients simultaneously received highly active antiretroviral therapy (HAART). Those patients with bulky tumors and very symptomatic at presentation received induction chemo before CRT. All cases were discussed in a multidisciplinary team (MDT). The choice of chemotherapy regimen was per the physician’s discretion.

After completing treatment, Complete Response (CR) was determined according to RECIST v1.1 from clinical, anorectoscopy and radiological images at 24 weeks. CR was defined as clinical (on anal inspection and examination), radiological (CT and MRI of the pelvis) and rectoscopy (no evidence of disease; suspected lesions were confirmed by a biopsy) disappearance of the disease. Abdominal perineal resection was performed in non-CR patients. Clinical follow-up included digital anal examination, anoscopy, and a clinical exam, monthly for 3 months, then every 3 months for the first two years and every 6 months for 3 to 5 years. A thorax CT and abdominal-pelvic MRI imaging were performed every 6 months during the follow-up period.

Forty out of the ninety-seven FFPE cases were selected based on quality and quantity of their purified DNA and RNA for further genomic and transcriptomic analysis. These 40 cases were represented by 22 patients with a complete clinical response and 18 patients with no complete clinical response after chemoradiotherapy.

### 2.3 Exome sequencing and bioinformatics analysis

Genomics DNA was isolated from FFPE samples using the “ReliaPrep™ FFPE gDNA Miniprep System” (Promega) according to the manufacturer’s instructions. The Nanodrop ND-1000 spectrophotometer (Thermo Scientific) was used to measure concentrations and the absorbance ratios at 260 nm and 280 nm for each sample, with the latter serving as an indicator of sample purity. All samples showed an acceptable 260/280 ratio value (greater than 1.78). The Qubit® 2.0 fluorometer (Thermo Scientific) was used to accurately measure the amount of DNA extracted from FFPE. Sample concentrations were determined using the “DNA BR Assay” kit (Thermo Scientific). The libraries were prepared using the Library Preparation Kit with Enzymatic Fragmentation 2.0 and the Exome Enrichment Kit 2.0 Plus (Twist Biosciences). The resultant libraries were also measured with the Qubit and DNA BR Assay kit (Thermo Scientific). The indexed libraries were circularized using the “Element Adept Library Compatibility v1.1” kit. Sequencing was performed with paired-end reads of 150 base pairs according to the recommendations provided by Element Biosciences AVITI™ platform at Helixio facility (Saint-Beauzire - France). The number of sequences ranged from 25 to 58 million. The GC content percentage was between 50% and 61%, and the duplication percentage also ranged from 13% to 30%. All these quality conditions have produced a validated dataset.

All short-read files from 40 patients (R1 and R2 fastq) were aligned against GRCh.38 with the BWA-MEM algorithm. A GATK standard pipeline, with Mutect2 caller, for “tumor-only” samples was applied. The mean coverage reached was 122x (ranging from 44 to 184x). The resulting raw VCFs were filtered to ensure that the variants obtained met the ‘PASS’ quality criteria and were of somatic origin. For annotation steps an up to date pipeline with SnpEff and dbNSFP resources were used. For tumor mutational burden (TMB) only non-synonymous variants (predicted with SnpEff as High or Moderate impact) were taken into account, and they also will have at least >= 10 reads for the alternative allele and also >= 5% for variant allele frequency (VAF). A driver discovery tool dNdScv was used to detect driver genes that could be under a positive selection in this cohort (17).

### 2.4 RNA sequencing and bioinformatics analysis

Total RNA was isolated from ASCC FFPE samples using the ReliaPrep™ FFPE Total RNA Miniprep System (Promega) following standard manufacturer’s protocol. RNA concentration and integrity were measured on an Agilent 2100 Bioanalyzer (Agilent Technologies). RNA samples with RNA integrity number (RIN) over 5 were considered for RNA sequencing. The RNA samples were processed for directional RNA-seq library construction using the NEBNext® rRNA Depletion v2 (Human/Mouse/Rat) module and the NEBNext® UltraTM II Directional RNA Library Prep kit (New England Biolabs) according to the manufacturer’s protocol. We performed 150 nt paired-end sequencing using an Element Biosciences AVITI™ platform at Helixio facility (Saint-Beauzire - France) and obtained ∼30 million clusters per sample with 92% >Q30. The RNAseq raw data has been submitted to the NCBI Gene Expression Omnibus database with the accession number GSE312258 (https://www.ncbi.nlm.nih.gov/geo/query/acc.cgi?acc=GSE312258). The raw short-read sequences were quality-checked and trimmed to remove adapters and low-quality bases using the Rfastp R/Bioconductor package. The preprocessed reads were then aligned and mapped to the human genome reference GRCh38 using the Subread aligner algorithm provided by the Rsubread R/Bioconductor package. The aligned reads (BAM files) from each sample were used to calculate gene expression abundance at the whole-genome level using the featureCounts function provided by the Rsubread package. To identify differentially expressed genes between NCR and CR ASCC, we computed fold changes and adjusted p-values using the edgeR R/Bioconductor package based on the normalized log2-based count per million values. Genes showing a log-fold change greater than 1 and an adjusted p-value below 0.05 were considered significantly differentially expressed. Functional enrichment analysis and Gene Set Enrichment Analysis (GSEA) of differentially expressed genes will be performed with the clusterProfiler R package. Tumor immune cell infiltration and T cell dysfunction/exclusion scores were estimated using five algorithms provided by the immunedeconv R/Bioconductor package (https://github.com/omnideconv/immunedeconv) on normalized count matrices. In addition, we used the Estimate Systems Immune Response (EaSIeR) R/Bioconductor package to characterize the tumor-immune microenvironment of ASCC based on RNAseq profiles (18). Immune response scores predicted by EaSIer were further compared among CR and NCR cases using the Rank Product test implemented in the RankProd R/Bioconductor package. Unsupervised hierarchical clustering analysis and heatmaps representations were performed with the MultiExperimentViewer (MeV 4.9.0) software.

### 2.5 HPV metatranscriptomic and p16 immunohistochemistry analyses

For metatranscriptomics, the obtained RNA-Seq data were processed using the bioBakery suite of tools: KneadData was used to separate the human and the nonhuman reads; taxonomic profiling was performed using MetaPhlAn to identify and quantify microbial taxa at species level present in the anal samples (19). Formalin-fixed and paraffin-embedded (FFPE) tissue sections were cut and reviewed by a specialist pathologist (JA) to confirm the presence of invasive ASCC. Freshly cut slides were stained for p16 as a surrogate marker of HPV infection, using a monoclonal anti-p16 antibody on an automated Leica Bond III IHC platform. Slides were categorized as p16 positive or negative by a specialized pathologist blinded to clinical outcomes. p16 was considered positive in case of a diffuse, nuclear and cytoplasmic, moderate to strong staining of tumor cells. Negative cases had < 10% tumor cells stained at any intensity.

### 2.6 Statistical analysis of clinicopathological and follow-up data

The Chi-square test was used to compare categorical data between groups, while Wilcoxon rank-sum test was used for continuous data. Kaplan-Meier curves and the log-rank test were used to analyze DFS and OS data. DFS was measured from the first day of CRT to clinical or radiological recurrence or death from any cause. OS was measured from treatment initiation to death from any cause, as previously reported (13). Two-tailed p-values were calculated and p-values < 0.05 were considered as significant.

## 3. RESULTS AND DISCUSSION

### 3.1 Patient cohort and treatment response

Ninety-seven FFPE samples of non-metastatic ASCC patients with treatment response and follow-up data after definitive chemoradiotherapy were recruited for further mutational and transcriptome based analysis. Clinical and demographic data are summarized in Table 1. Eighty-two patients were treated with mitomycin-5-FU (85%), 5 patients received cisplatin-5-FU (5%), and 10 patients received mitomycin-capecitabine (10%), concomitantly with 3D-pelvic radiotherapy. Sixty-nine patients reported complete response (CR=71%) at 6 months after initiation of CRT, while partial or stable response was reported in 28 patients (NCR=29%). No significant differences were found between CRT regimens according to CR rate and follow-up (p>0.05). Tumor stage and lymph node status were associated with treatment response (p=0.008 and p=0.043, respectively) and disease-free survival (p<0.0001 and p=0.0083, respectively).

**Table 1.**
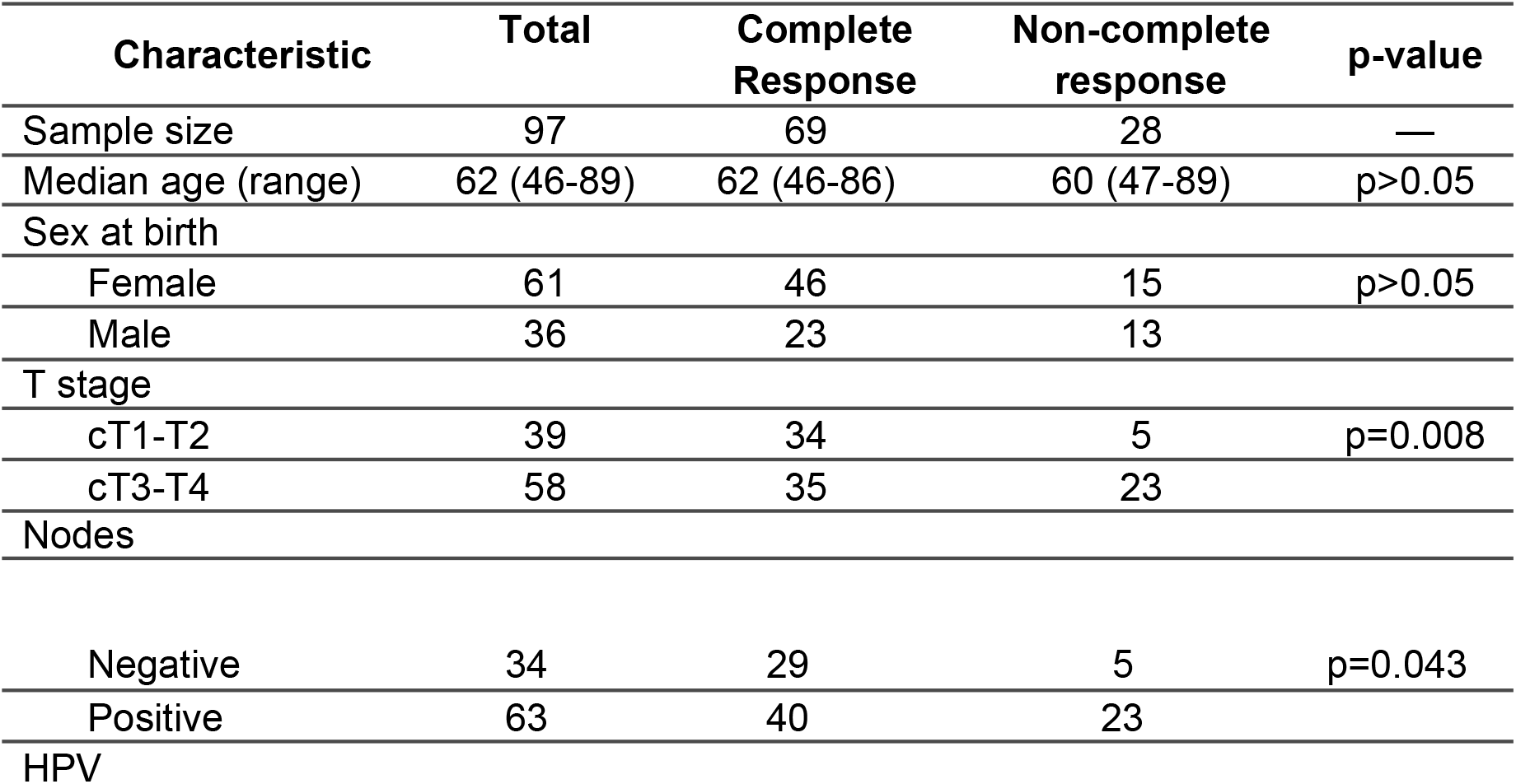
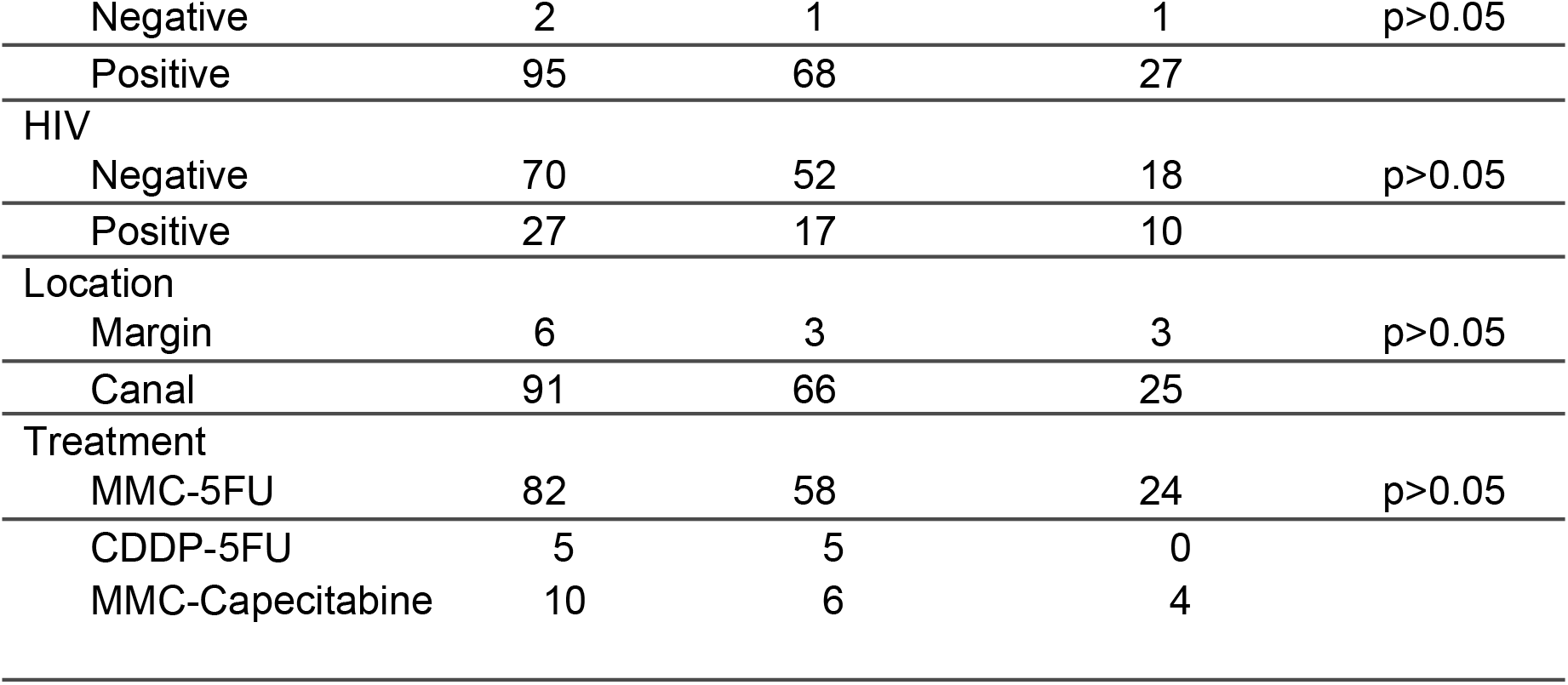
Clinicopathological characteristics of the ASCC cohort.

### 3.2 HPV infection among CRT responder and non-responder ASCC

Ninety-eight percent of the ASCC (95 out of 97) were HPV-positive cases according to p16 IHC analysis (Table 1). Virome analysis of the 40 ASCC samples profiled by RNAseq showed that 36 out of 38 HPV-positive cases were infected by high-risk oncogenic subtypes (Figure 1). Consistent with p16 IHC results (38/40), Alphapapillomavirus-9 (comprising genotypes HPV16, 31, 33, 52, and 58) was detectable in the majority of samples (33/40). Additionally, Alphapapillomavirus-7 (HPV18, 39, 59, 68, 45, 70), Alphapapillomavirus-5 (HPV26, 51, 69, 82), and Alphapapillomavirus-10, which includes the low-risk genotypes HPV6 and HPV11, were also detected in a subset of ASCC. Non-significant associations with CRT response or other clinicopathological variables were detected for HPV infection as determined by p16 or RNAseq virome analysis (p>0.05).

**Figure 1.**
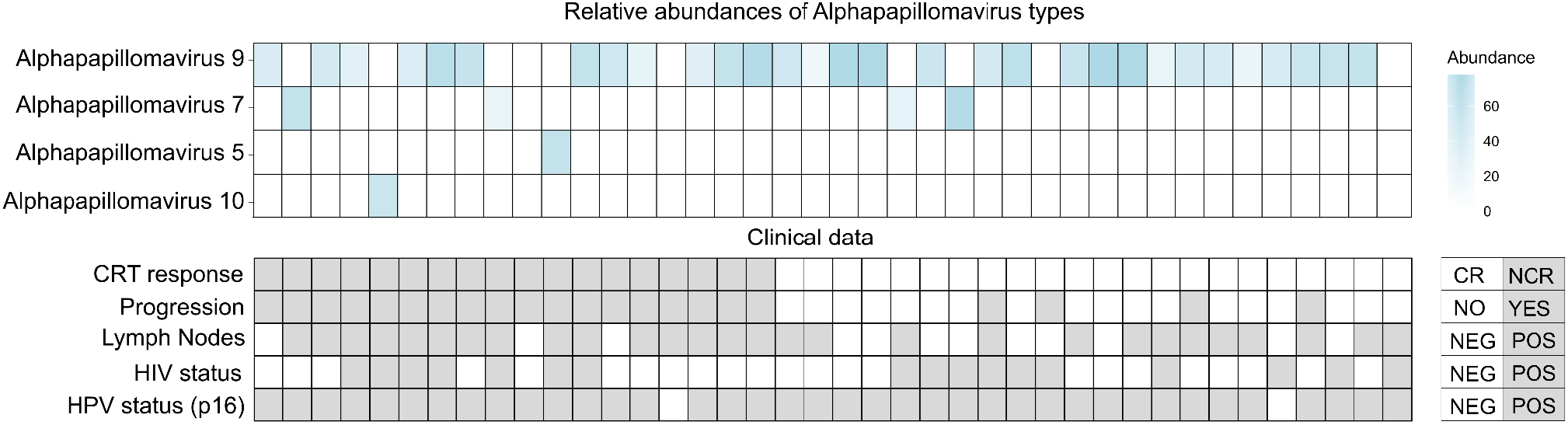
Heatmap of relative abundances of Alphapapillomavirus species detected in 40 ASCC samples, as assessed by metatranscriptomic analysis. Below the heatmap, a tile plot aligns samples with their clinicopathological variables including CRT response, progression, lymph node status and HIV and HPV status based on p16 IHC.

### 3.3 Transcriptome profile of responder and non-responder ASCC

Statistical analysis of RNA-Seq data revealed 350 differentially expressed genes (DEGs) between CR and NCR cases (p-value<0.01; FC>2), 87% were coding RNAs and 13% were ncRNAs. Among the deregulated genes, 261 were up-modulated and 89 down-modulated genes in CR compared with NCR cases (Figure 2A and Supplementary Table S1). Functional enrichment analysis of 350 DEGs revealed specific functional bioprocess strongly related to adaptive immunity, epidermis development, cell differentiation, cytokine production (Figure 2B) and signaling pathways associated with TNFA/NFkB, epithelial to mesenchymal transition, KRAS and IL6-JAK-STAT3 signaling (Supplementary Figure 1). Interestingly, we identify several multifaceted tumor suppressor related genes involved with the modulation of the TP53 pathway and the immune response among the most significant CR up-modulated genes compared to NCR cases such as *FDCSP, ALDOB, ADGRB1* and *SPINK7* (Figure 2C-D, Supplementary Table 1). Several CR up-modulated genes were significantly associated with longer DFS and OS outcomes (Figure 2D, Supplementary Table 1).

**Figure 2.**
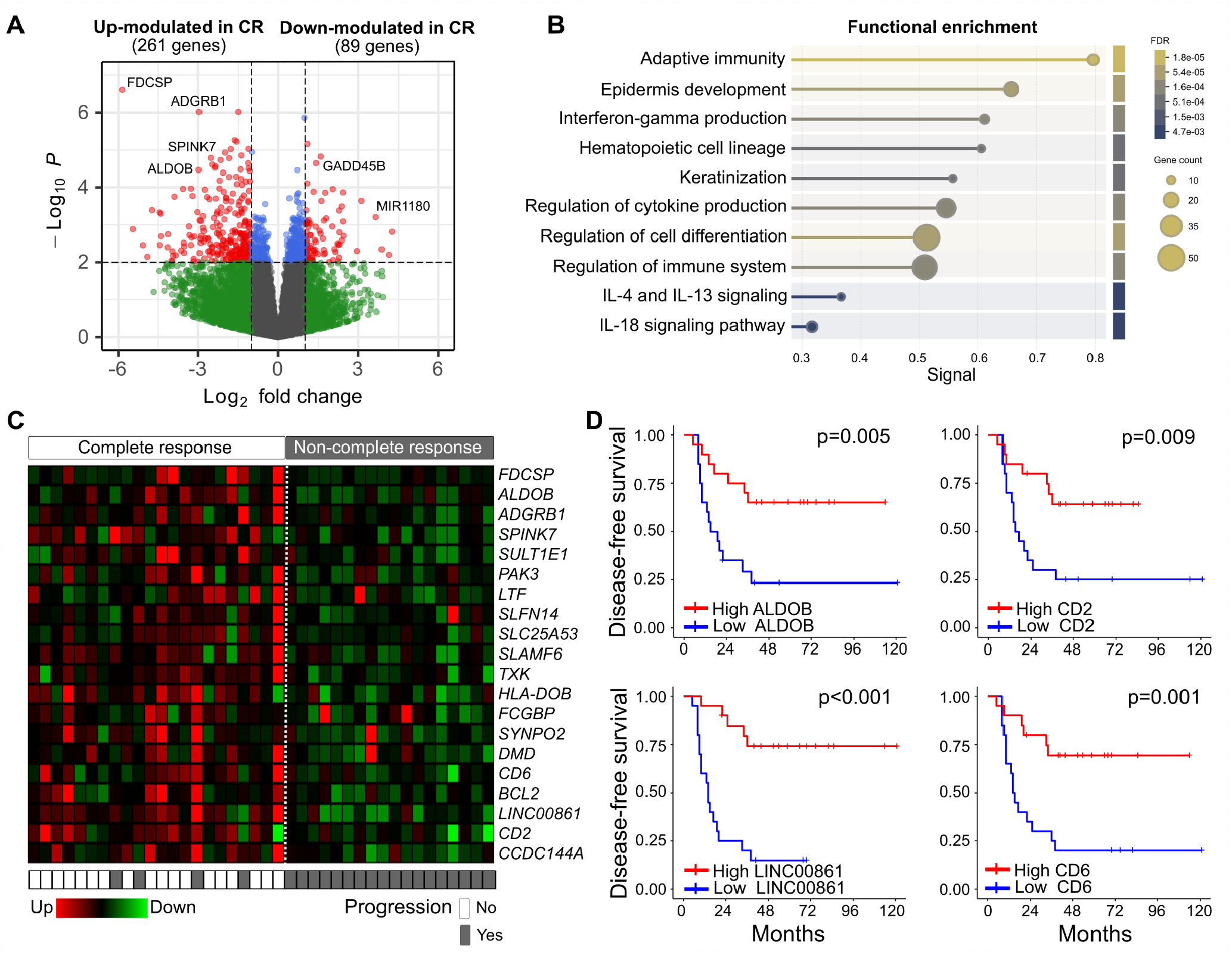
Transcriptome changes among 40 non-metastatic ASCC according to CRT treatment response. **A)** Volcano plot of 350 differentially expressed genes among CR and NCR ASCC patients (p<0.01; FC>2). **B)** Functional enrichment analysis of the differentially expressed genes based on Gene Ontology Biological process, KEGG and Reactome pathway databases (30). **C)** Heatmap of top 20 most significantly deregulated genes in CR cases. **D)** Representative Kaplan-Meier curves of a coding (ALDOB), a non-coding (LINC00861), and two immune-related genes (CD2 and CD6) up-modulated in CR cases and associated with ASCC disease-free survival.

Among the most DEGs, the *Follicular dendritic cell secreted protein* (FDCSP) has recently emerged as a significant biomarker in various cancer contexts, particularly in relation to immune responses and cancer prognosis. FDCSP is highly expressed in HPV-positive head and neck squamous carcinoma (HNSC) and is associated with a favorable prognosis. Its expression correlates with increased infiltration of T follicular helper cells, which are crucial for effective immune responses. FDCSP’s function is linked to chemokine pathways, particularly CXCL13, suggesting its role in modulating immune responses in HPV+ HNSC (20).

As described, several of the most significantly CR up-modulated genes act as tumor suppressors by regulating key pathways involved in cancer progression. Aldolase B (ALDOB), a glycolytic enzyme, suppresses tumor growth in hepatocellular and gastric cancers by inhibiting the Akt pathway and modulating the immune microenvironment (21,22). ALDOB downregulation leads to increased TGF-β, immune evasion, and impaired CD8+ T cell function (23). Adhesion G protein-coupled receptors B1 (ADGRB1) prevents p53 degradation by inhibiting Mdm2, maintaining p53-mediated tumor suppression; its loss results in lower p53 levels and enhanced tumor proliferation (24,25). SPINK7 (also known as ECRG2) encodes a serine protease inhibitor and p53 target, limits cancer cell proliferation, migration, and invasion; its absence is linked to chemoresistance and increased malignancy including squamous esophageal and oral squamous cell carcinoma (26). Loss of SPINK7 expression can lead to resistance against DNA-damaging anticancer drugs, highlighting its potential as a therapeutic target (27). NLRC3 functions as a negative regulator of signaling pathways activated by Toll-like receptors (TLRs) and the DNA sensor STING in response to viral infections (28). NLRC3 associates with PI3Ks, inhibiting the activation of the PI3K-dependent kinase AKT following the binding of growth factor receptors or TLR4. These findings underscore NLRC3 as an inhibitor of the mTOR pathway, an immune regulator, and a tumor suppressor gene (28,29).

This study showed that *FDCSP, ALDOB, ADGRB1, SPINK7* genes, and others shown in Supplementary Table S1, are deregulated in NM-ASCC in association with clinical response to CRT treatment. Importantly, a recent study demonstrates that activation of inflammatory pathways such as IFNγ, IFNα, TNFα signaling via NF-κB, and EMT were significantly enriched in ASCCB, and EMT were significantly enriched in ASCC tumors that respond poorly to chemoradiotherapy (31). Elevated expression of interferon-induced transmembrane protein 1 (IFITM1), increased regulatory T-cells, and higher levels of the chemokine CXCL2 in blood were also associated with reduced freedom from locoregional failure and distant metastasis after CRT (31).

Further studies need to be performed among independent cohorts to corroborate the relevance of these transcripts as prognostic and predictive biomarkers and its application in clinical settings, particularly in enhancing the effectiveness of CRT in NM-ASCC patients.

### 3.4 Immune infiltrate populations within distinct tumor treatment response

Given the aforementioned strong association of the transcriptome changes with immune-related themes, we performed a computational evaluation of tumor immune infiltrate among CR and NCR cases using CIBERSORT, xCell, MCPcounter, Quantiseq and TIMER algorithms. Supporting the GO and functional annotation observations, tumor immune infiltrate analysis based on RNA-seq data identified the enrichment of different subpopulations of T and B-cells in CR cases compared to NCR (Figure 3A). The CD8+ central memory T cells was the most significantly enriched immune type among CR cases (p=0.008) associated with longer and DFS (p=0.005) and OS (p=0.003) (Figure 3C, D). The CD4+ memory resting B cells were also enriched in CR cases compared to NCR (p=0.01) in association with longer DFS (p=0.021) and OS (p=0.002). The remaining immune cell infiltrates enriched in CR cases were not significantly associated with outcomes (p>0.05). Interestingly, T cell CD4+ Th1 and Macrophage M1 were significantly depleted among CR compared with NCR cases (p=0.044 and p=0.01) (Figure 3A). In addition, T cell CD4+ memory resting (p=0.041), Tregs (p=0.008) and myeloid dendritic cell activated (p=0.011) were significantly depleted among HIV-positive compared with HIV-negative cases (Figure 3B).

**Figure 3.**
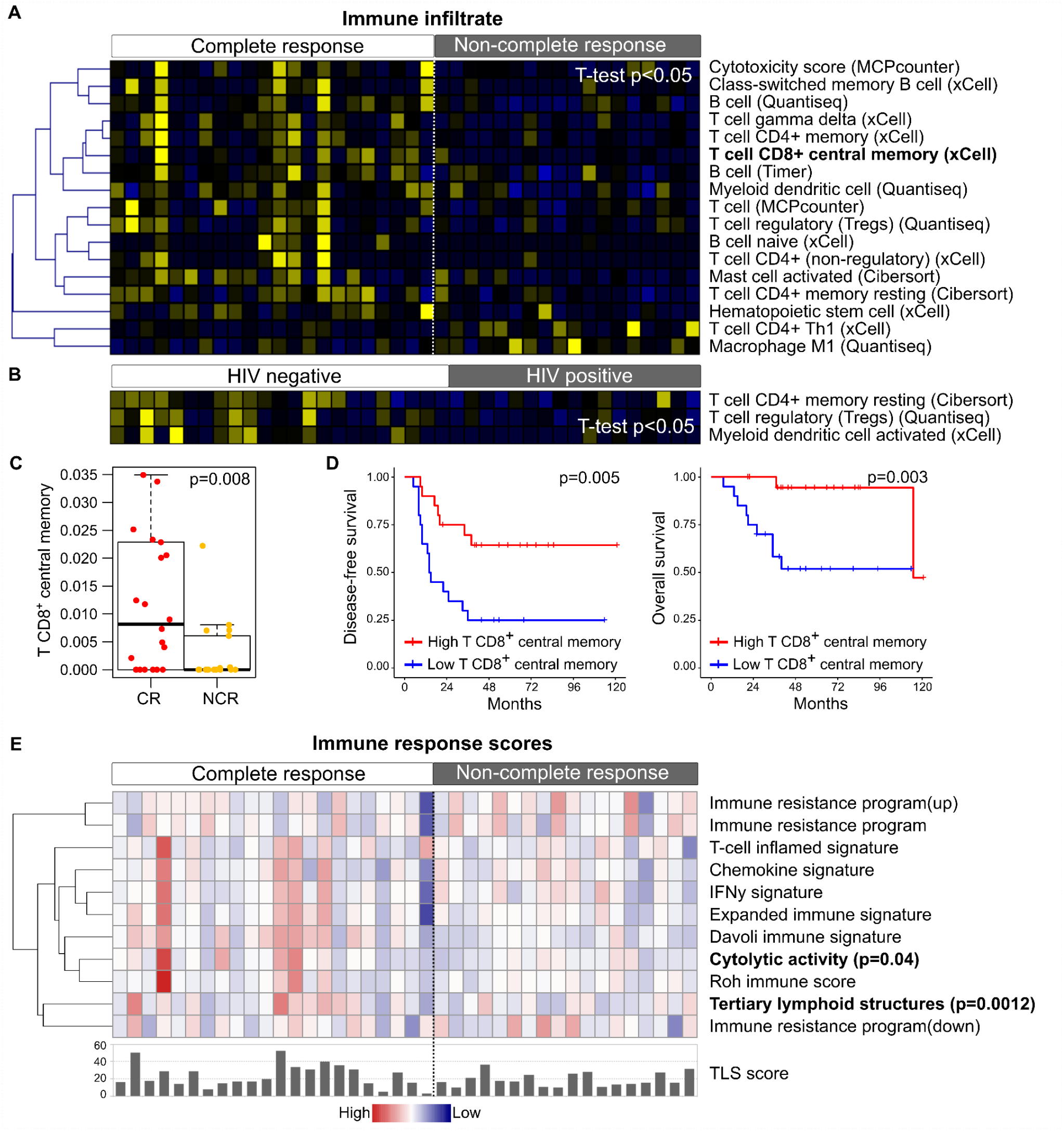
Transcriptome-based predictors of the tumor immune infiltrate and immune response among ASCC patients according to the clinical response to treatment. **A)** Tumor immune infiltrate fractions (TILs) differentially enriched among responder and non-responder patients (p<0.05) as estimated by CIBERSORT, xCell, MCPcounter, Quantiseq and TIMER algorithms. **B)** TILs differentially enriched among HIV-negative and HIP-positive cases (p<0.05). **C)** Boxplot of T cell CD8+ central memory tumor infiltrates among patients with CR and NCR response (p=0.008). **D)** Survival analysis of the T cell CD8+ central memory tumor infiltrates among NM-ASCC patients. **E)** Immune response scores as estimated by the EaSIeR R/Bioconductor package. Increased tertiary lymphoid structure (TLS) and cytolytic activity were significantly detected among CR cases compared to NCR (p=0.0012 and p=0.05, respectively).

We further employed the Estimate Systems Immune Response (EaSIeR) tool to generate a high-level representation of the anti-tumor immune responses in the tumor microenvironment of CR and NCR cases. Briefly EaSIer computes immune response scores based on gene expression signatures including immune cytolytic activity, chemokine, IFNy, T-cell inflamed, immune resistance program and tertiary lymphoid structures (TLS) signatures among others. Interestingly, the TLS (FC=4.4; p=0.0012) and cytolytic activity (FC=1.7; p=0.04) scores were significantly increased among CR compared with NCR cases (Figure 3D). The immune cytolytic activity score represents the level of two cytolytic effectors, granzyme A and perforin, which are overexpressed upon CD8+ T cell activation. The TLS score is derived from differentially expressed genes in tumors with TLS.

The enrichment of B cells, T cells, particularly T central memory cells, and dendritic cells among CR cases could be explained by TLS associated with CR cases. TLS are privileged sites of lymphoid neogenesis within tumors for the recruitment and activation of central-memory T and B cells that circulate and limit cancer progression (32). TLS has been associated with a favorable prognosis in various cancers such as non-small cell lung cancer, colorectal, gastric, pancreatic, and esophageal cancer among others (33). Recently, Wang et al. showed that TLS predicts the response to neoadjuvant therapy and recurrence-free survival of patients with locally advanced rectal cancer(34).

Overall, these findings suggest that CR cases had a significant enrichment of TLS with T CD8+ central memory cells that could facilitate an increased CD8+ regional memory T cell in CR compared to NCR ASCC cases. CD8+ regional memory T cells have been consistently associated with favorable prognosis in multiple cancer types, including lung cancer, endometrial adenocarcinoma, bladder urothelial carcinoma, cervical cancer, and gastric cancer (35,36). High densities of these cells within tumors correlate with improved survival rates and better clinical outcomes (37–39). Further studies need to be performed to corroborate the CD103+CD8+ TILs among NM-ASCC in association with CRT response.

### 3.5 Mutational profile of responder and non-responder ASCC

Exome-seq was performed in pretreatment biopsies from 40 patients using the Exome Enrichment Kit 2.0 Plus (Twist Biosciences), The mean coverage for all samples was 122.4X (min 43.9X and max 184.6X) that allow the identification of 172 somatic mutations across 21 cancer driver genes. Missense variants accounted for 70% of the detected mutations, while nonsense mutations represented 14%. Frameshift mutations and in-frame indels each constituted approximately 2% and 14% of the total, respectively. We combined a bioinformatics approach, using the dNdScv algorithm, with a thorough literature review to identify cancer driver genes (40). In this sense, we selected genes with significant driver mutations (q global <0.1) and those mutated genes previously associated with ASCC. Among these, we detected mutations affecting SLAMF7 (65%), GOLGA6L9, ZNF208 and ZNF429 (43%), RBM38 (40%), ZNF430 (35%) and MTCH2 (32.5%) and for the genes that was previously associated to ASCC we found that KMT2C (18%), KMT2D (13%), PIK3CA, FBXW7, ATM, RB1 and PTEN account for (10%) and finally EP300, BRCA2, APC, CDKN2A (8%) and JAK2, NOTCH1 and, TP53 (5%) (Figure 4A). The mean Tumor Mutational Burden (TMB) for all samples was 6 mut/Mb ranging from 2.6 to 18.2. Patients with a higher number of mutations per megabase showed a trend towards complete response to treatment, although this difference was not statistically significant (p=0.07) (Figure 4B). In addition, patients with high TMB showed a shorter DFS (p=0.027) and OS (p=0.03) compared with low TMB cases (Figure 4C).

**Figure 4.**
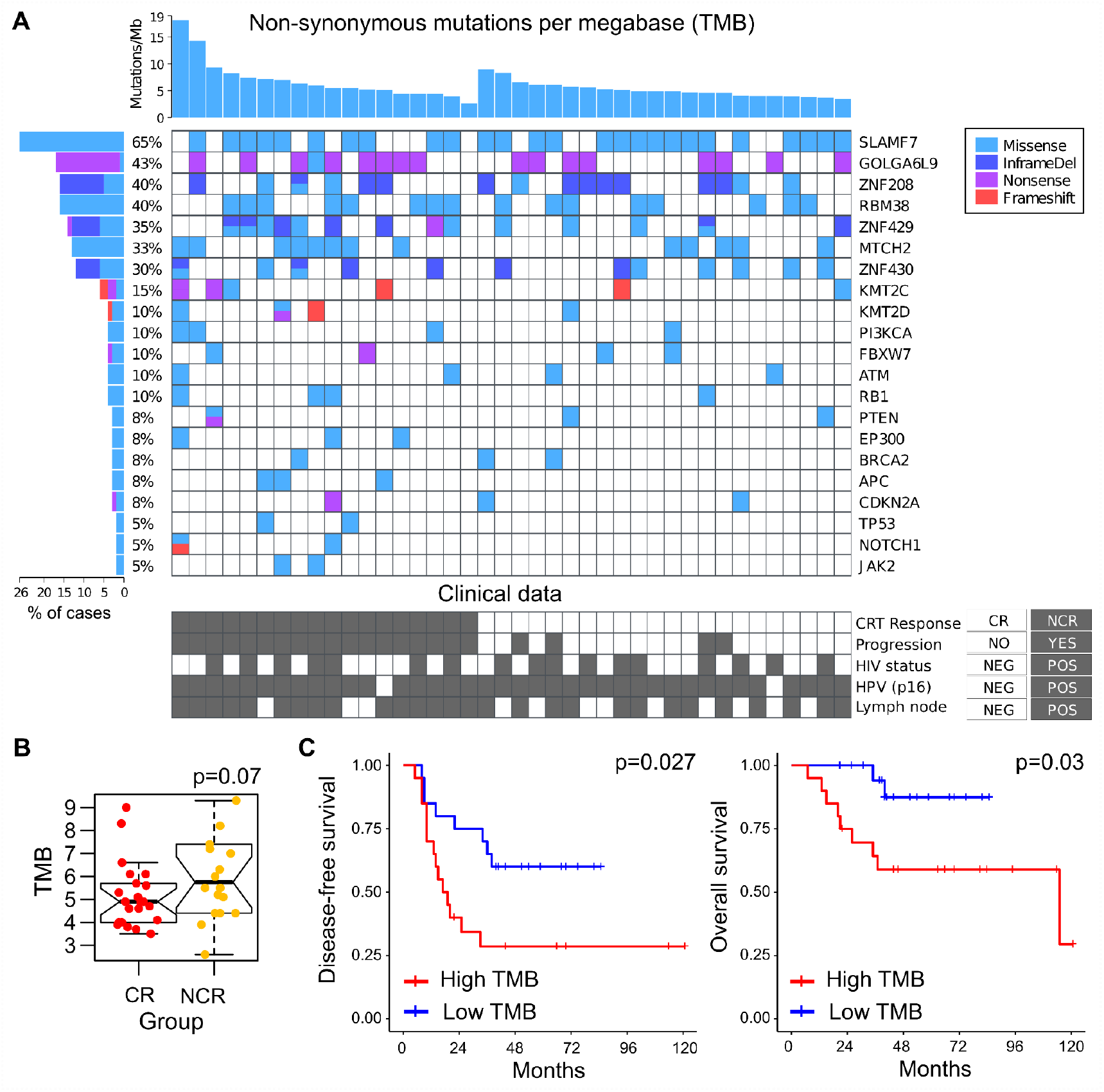
Mutational profile of NM-ASCC according to the CRT response. **A)** Tile plot showing recurrent altered cancer driver genes and the significant genes that were under positive selection based on dNdScv algorithm among 40 non-metastatic ASCC. Non-significant associations with CRT response were detected for any of the mutational variants identified (p>0.05). **B)** Tumor Mutational Burden (TMB) among patients with complete (CR) and non-complete (NCR) response to CRT. **C)** Survival analysis of the TMB among NM-ASCC patients. Tumors with high TMB were associated with shorter DFS (p=0.027) and OS (p=0.03) compared with low TMB tumors.

High TMB is often regarded as a marker of increased immunogenicity and a predictor of response to immune checkpoint inhibitors in various cancers. However, in patients receiving standard treatments such as chemoradiotherapy (CRT), tumors with high TMB may carry numerous driver mutations that contribute to more aggressive tumor biology and greater resistance to DNA-damaging therapies (41). Non-significant associations were detected for any of the mutational variants identified with the CRT response (p>0.05).

### Study limitations

Despite our comprehensive analysis of clinical, mutational, and transcriptomic characteristics, the data on treatment and clinical responses are derived from a single-center cohort, which may not fully represent the variability found across different clinical environments. Moreover, the retrospective design introduces potential biases, including variability in treatment regimens, treatment adherence, and patient comorbidities, which could influence the results. Additionally, while exome sequencing and transcriptome analysis are robust methods, the absence of validation of some identified biomarkers and genes in independent cohorts is a significant limitation. The findings should be viewed as preliminary until confirmed in larger, more diverse studies. While we observed interesting associations between the transcriptomic profile and treatment response, the limitations in immune infiltrate analysis, particularly in identifying specific T and B cell subpopulations, require further investigation. Additional methods, such as a more detailed characterization of the tumor microenvironment, could enhance our understanding of the mechanisms driving treatment response in ASCC.

## 4. Conclusions

Comprehensive characterization of non-metastatic ASCC at mutational, transcriptomic and immune levels allowed us to identify the most relevant changes in the context of CRT response and survival outcomes. A comparison of the mutational profile identified in this non-metastatic cohort revealed novel putative cancer driver genes frequently altered in ASCC such as SLAMF7 and GOLGA6L9 and previously reported genes but not associated with clinical response to CRT treatment. Our study highlights key molecular and immune markers that could improve the clinical management of ASCC patients. We identified a gene expression signature expressed in CR cases (e.g. FDCSP, ALDOB, ADGRB1, SPINK7) and downregulated in NCR, which are associated with good prognosis and that may serve as potential biomarkers of CRT response. Tumor-immune infiltrate analysis revealed that responders to CRT exhibited enrichment of T and B cell subpopulations, particularly CD8+ central memory T cells and CD4+ resting memory B cells, which correlated with improved survival outcomes and the presence of tertiary lymphoid structures likely plays a role in this immune enrichment environment. Together, these findings underscore the potential of integrating molecular and immune markers into clinical practice to better predict treatment response and guide personalized therapies for ASCC patients. Further validation in independent cohorts is necessary to confirm the clinical relevance of these biomarkers and their application in therapeutic decision-making.

## Data Availability

All data produced in the present study are available upon reasonable request to the authors

## Author Contributions

All the authors have directly participated in the preparation of this manuscript and have read and approved the final version submitted and declare no ethical conflicts of interest. SI and MCA conceived the study, performed formal analysis, and wrote the article. GM, EL, DP, SB, CL, NB, ER and JA were responsible for methodology, research assistance, genomics data analysis, and clinical data curation of participants.

## Informed consent and patient details

This retrospective study was approved by the Hospital Paris Saint Joseph (HPSJ) ethics committee. All patients provided informed consent for their data collection according to the recommendation of the ethics committee and in accordance with the European Union General Data Protection Regulation (GDPR).

## Acknowledgments

We thank the patients who participated in this research and their relatives for their time, altruism, and generosity. We extend our heartfelt gratitude to Dr. Esteban Cvitkovic for his invaluable contributions and unwavering support to this study and previous research efforts in rare cancer malignancies.

