## Supplementary Table 1 for "Transcriptomic Immune-related Signature Predictive of Chemoradiotherapy Response in Anal Squamous Cell Carcinoma"

**Supplementary Table 1.** Most significantly up-modulated transcripts in complete response ASCC cases

| Gene | Description | Fold Change | p-value adjusted | DFS p-value | DFS HR |
|---|---|---|---|---|---|
| *FDCSP* | Follicular dendritic cell secreted protein | 57.8 | 0.005 | 0.573 | — |
| *ALDOB* | Aldolase, fructose-bisphosphate B | 7.9 | 0.030 | 0.005 | 0.29 |
| *ADGRB1* | Adhesion G protein-coupled receptor B1 | 7.8 | 0.007 | 0.932 | — |
| *SPINK7* | Serine peptidase inhibitor Kazal type 7 | 5.8 | 0.024 | 0.234 | — |
| *SULT1E1* | Sulfotransferase family 1E member 1 | 5.6 | 0.028 | 0.054 | — |
| *PAK3* | p21 (RAC1) activated kinase 3 | 5.2 | 0.030 | 0.161 | — |
| *LTF* | Lactotransferrin | 5.1 | 0.030 | 0.173 | — |
| *SLFN14* | Schlafen family member 14 | 4.6 | 0.047 | 0.023 | 0.37 |
| *SLC25A53* | Solute carrier family 25 member 53 | 4.5 | 0.025 | 0.144 | — |
| *SLAMF6* | SLAM family member 6 | 4.2 | 0.033 | 0.1 | — |
| *TXK* | TXK tyrosine kinase | 4.0 | 0.024 | 0.006 | 0.30 |
| *HLA-DOB* | MHC class II, DO beta | 3.6 | 0.040 | 0.044 | 0.41 |
| *FCGBP* | Fc fragment of IgG binding protein | 3.6 | 0.024 | 0.152 | — |
| *SYNPO2* | Synaptopodin 2 | 3.4 | 0.023 | 0.007 | 0.30 |
| *DMD* | Dystrophin | 3.1 | 0.022 | 0.017 | 0.35 |
| *CD6* | CD6 molecule | 3.0 | 0.040 | 0.001 | 0.23 |
| *BCL2* | BCL2 apoptosis regulator | 2.9 | 0.022 | 0.443 | — |
| *LINC00861* | Long intergenic non-coding RNA 861 | 2.8 | 0.007 | 2.33E-05 | 0.13 |
| *CD2* | CD2 molecule | 2.7 | 0.031 | 0.009 | 0.31 |
| *CCDC144A* | Coiled-coil domain containing 144A | 2.6 | 0.044 | 0.289 | — |
| *RNU5A-1* | RNA, U5A small nuclear 1 | 2.6 | 0.024 | 0.004 | 0.28 |
| *PTPN22* | Protein tyrosine phosphatase non-receptor type 22 | 2.3 | 0.047 | 0.335 | — |
| *NLRC3* | NLR family CARD domain containing 3 | 2.1 | 0.027 | 0.002 | 0.25 |
| *KCNA3* | Potassium voltage-gated channel subfamily A member 3 | 2.1 | 0.023 | 0.001 | 0.15 |
| *P2RY8* | P2Y receptor family member 8 | 2.1 | 0.03 | 0.030 | 0.38 |
