## Supplementary figures and images for "Transcriptomic Immune-related Signature Predictive of Chemoradiotherapy Response in Anal Squamous Cell Carcinoma"

### Supplementary Figure 1

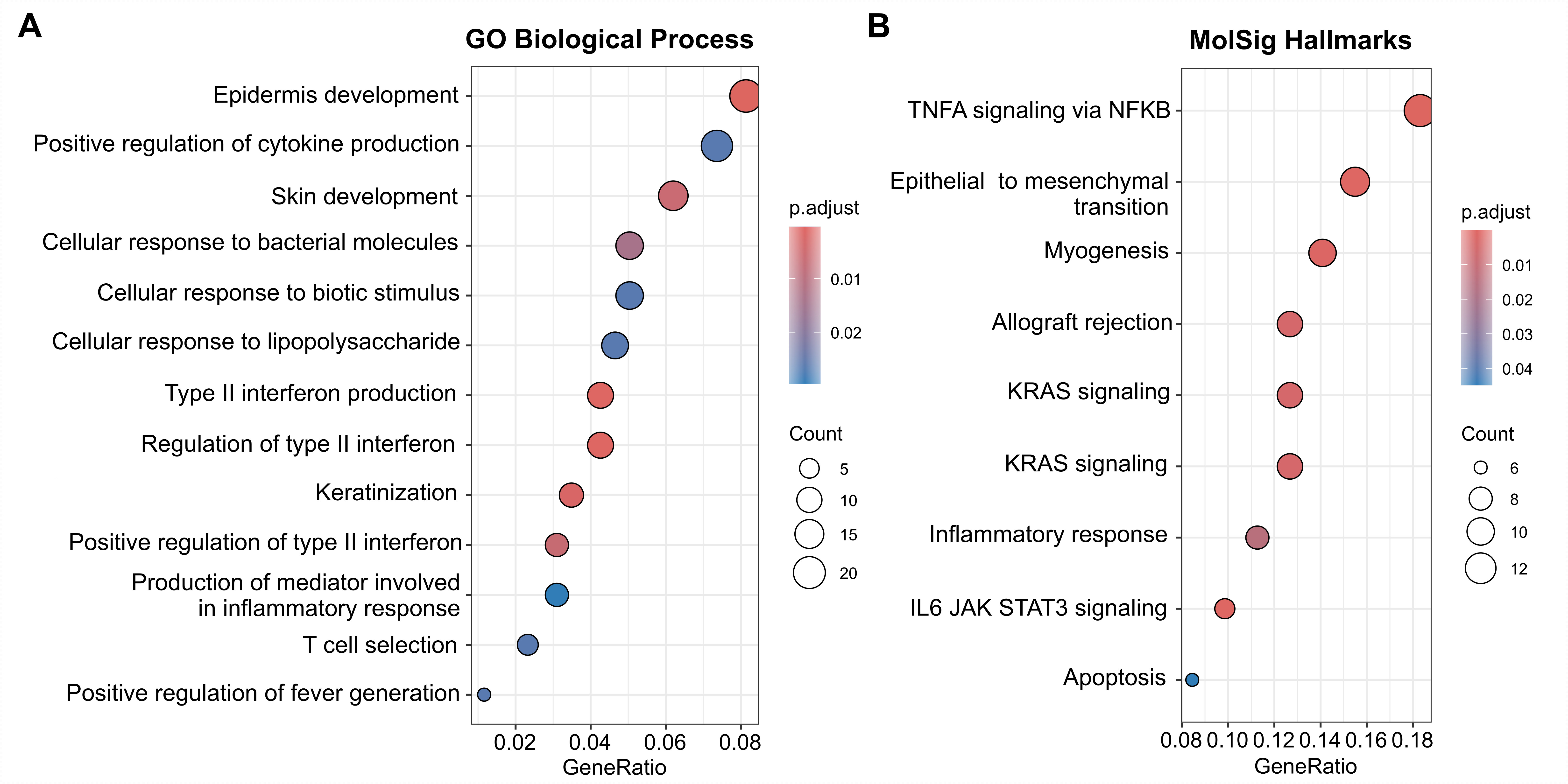
